# Changes in live births, preterm birth, low birth weight, and cesarean deliveries in the United States during the SARS-CoV-2 pandemic

**DOI:** 10.1101/2021.03.20.21253990

**Authors:** Alison Gemmill, Joan A. Casey, Ralph Catalano, Deborah Karasek, Tim Bruckner

## Abstract

**Background:** The SARS-CoV-2 pandemic and associated social, economic, and clinical disruption have been widely speculated to affect pregnancy decision-making and outcomes. While a few US-based studies have examined subnational changes in fertility, preterm birth, and stillbirth, there remains limited knowledge of how the pandemic impacted childbearing and a broader set of perinatal health indicators at the national-level throughout 2020. Here, we use recently released national-level data to fill this gap. Importantly, we, unlike earlier work, use time-series methods to account for strong temporal patterning (e.g., seasonality, trend) that could otherwise lead to spurious findings.

**Methods:** For the years 2015 to 2020, we obtained national monthly counts of births and rates (per 100 births) for six perinatal indicators: preterm birth (<37 weeks gestation), early preterm birth (<34 weeks gestation), late preterm birth (34-36 weeks gestation), low birth weight birth (<2500 g), very low birth weight birth (<1500 g), and cesarean delivery. We use an interrupted time-series approach to compare the outcomes observed after the pandemic began (March 2020) to those expected had the pandemic not occurred.

**Results:** For total births as well as five of the six indicators (i.e., all but the rate of cesarean delivery), observed values fall well below expected levels (p<.0001 for each test) during the entire pandemic period. Declines in preterm birth and low birth weight were largest in magnitude in both early and later stages of the 2020 pandemic, while those for live births occurred at the end of the year.

**Discussion:** Our findings provide some of the first national evidence of substantial reductions in live births and adverse perinatal outcomes during the SARS-CoV-2 pandemic. Only cesarean delivery appeared unaffected. These declines were not uniform across the pandemic, suggesting that several mechanisms, which require further study, may explain these patterns.

## Background

The SARS-CoV-2 pandemic and associated social, economic, and clinical disruption have been widely speculated to affect pregnancy decision-making and outcomes. While a few US-based studies have examined subnational changes in fertility, preterm birth, and stillbirth,^1-3^ there remains limited knowledge of how the pandemic impacted childbearing and a broader set of perinatal health indicators at the national-level throughout 2020. Here, we use recently released national-level data to fill this gap. Importantly, we—unlike earlier work—use time-series methods to account for strong temporal patterning (e.g., seasonality, trend) that could otherwise lead to spurious findings.

## Methods

For the years 2015 to 2020, we obtained national monthly counts of births and rates (per 100 births) for six perinatal indicators: preterm birth (<37 weeks gestation), early preterm birth (<34 weeks gestation), late preterm birth (34-36 weeks gestation), low birth weight birth (<2500 g), very low birth weight birth (<1500 g), and cesarean delivery.

Data from 2020 were obtained from provisional estimates released by the National Center for Health Statistics in March 2021.^4^ As of March 17, 2021, these estimates were based on 99.87% of births occurring to US residents within the 50 states and the District of Columbia in 2020; disaggregated estimates were not available. Monthly counts of births and rates for the six perinatal indicators from 2015 to 2019 were obtained from the Centers for Disease Control and Prevention Wonder.^5^

We use an interrupted time-series approach to compare the outcomes observed after the pandemic began (March 2020) to those expected had the pandemic not occurred. We estimate expected values from March 2020 through December 2020 using well-established ARIMA routines (or Box-Jenkins methods).^6^ For the entire pandemic period, as well as for each month, we test whether observed values were significantly different from expected. A detailed description of these analyses appears in the Supplement. Institutional review board approval was not required because the deidentified data are publicly available.

## Results

Table 1 shows differences between observed and expected values for total births and the six perinatal indicators for the months of March through December 2020. Figures 1-7 plot monthly trends in expected and observed counts (for live births) and rates (for perinatal indicators) from 2015-2020.

**Table 1.** Difference between observed and expected values for live births (counts) and perinatal indicators (rates per 100 live births), March through December, 2020.

|  | Live births | Preterm birth | Late Preterm birth | Early Preterm Birth | Low birth weight | Very low birth weight | Caesarean Delivery |
| --- | --- | --- | --- | --- | --- | --- | --- |
| March | 1,055 | -0.64 * | -0.44 * | -0.18 * | -0.23 * | -0.08 | -0.06 |
| April | -5,098 | -0.53 * | -0.40 * | -0.06 | -0.09 | -0.04 | 0.05 |
| May | -10,619 * | -0.33 * | -0.24 * | -0.06 | -0.16 * | 0.01 | -0.03 |
| June | 1,668 | -0.45 * | -0.25 * | -0.12 * | -0.20 * | -0.05 | 0.37 |
| July | -7,235 | -0.29 * | -0.16 | -0.08 | -0.06 | -0.03 | 0.28 |
| August | -16,853 * | -0.17 | -0.07 | -0.09 * | -0.14 | -0.06 * | 0.00 |
| September | -9,753 * | -0.23 * | -0.15 | -0.09 * | -0.14 | -0.09 * | 0.29 |
| October | -15,812 * | -0.12 | -0.06 | -0.06 | -0.04 | -0.07 * | 0.32 |
| November | -12,952 * | -0.61 * | -0.30 * | -0.26 * | -0.36 * | -0.15 * | 0.18 |
| December | -19,624 * | -0.40 * | -0.32 * | -0.05 | -0.24 * | -0.04 | 0.07 |

\*p<.05

**Figure 1.**
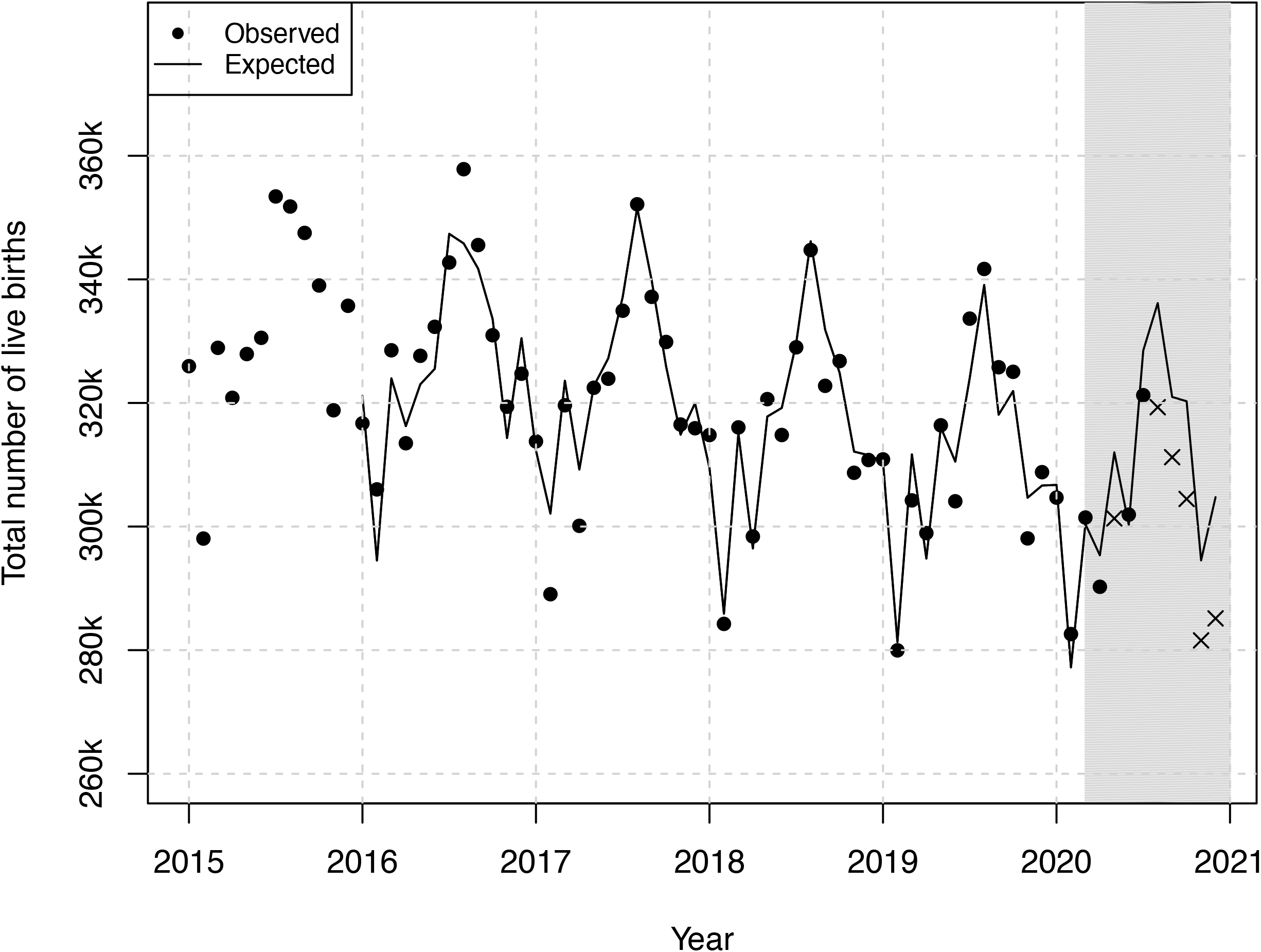
Observed and expected monthly trend of counts of live births, 2015-2020. Shaded area represents pandemic months (March through December 2020). Observed points marked with an X show values significantly different than expected during pandemic period. Includes 72 months beginning January 2015 and ending December 2020. Expected values were generated from a time series model using observed rates from January 2015 through February 2020. The first 12 months were lost to modeling.

**Figure 2.**
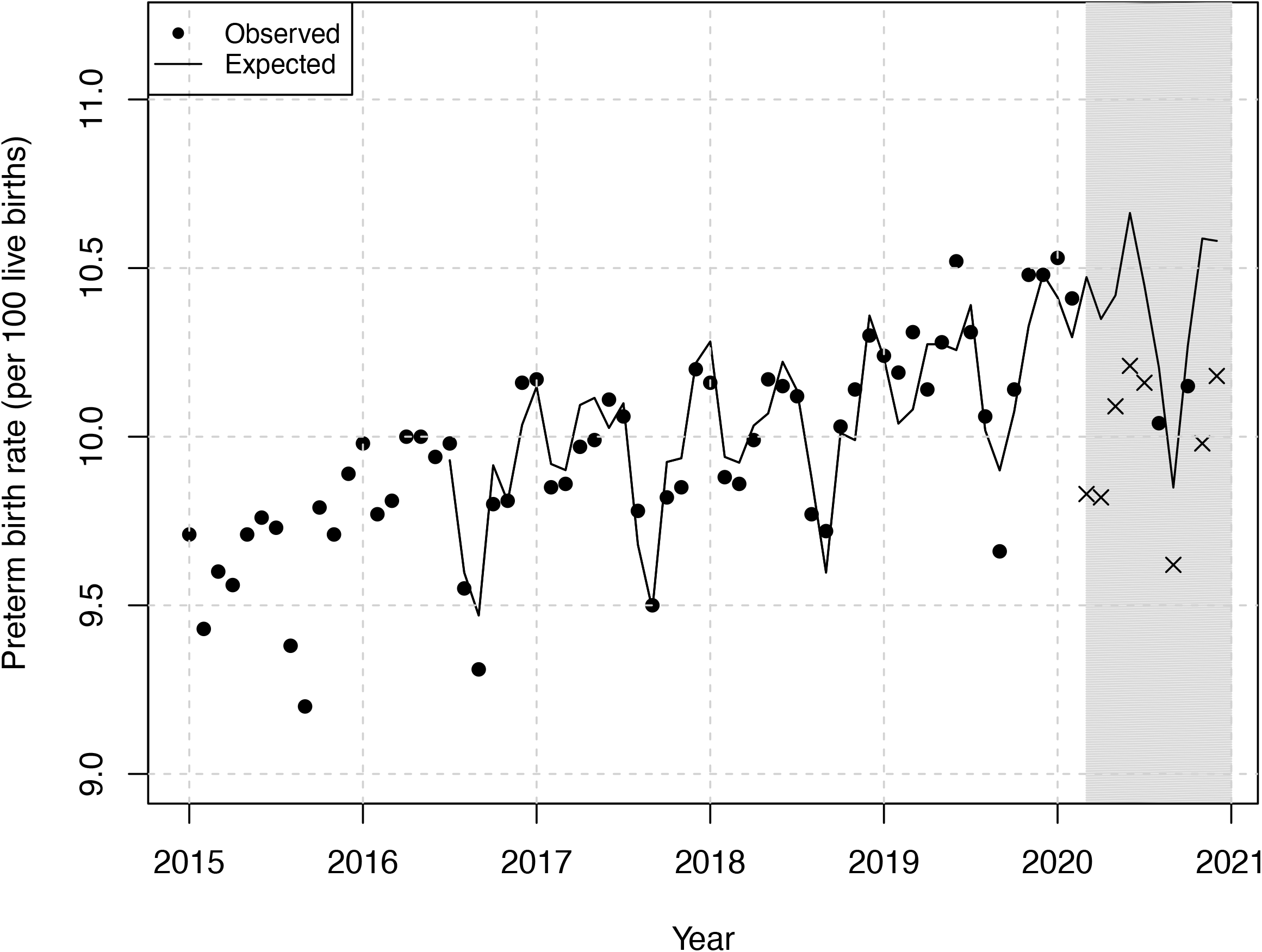
Observed and expected monthly trend of the preterm birth rate (per 100 live births), 2015-2020. Shaded area represents pandemic months (March through December 2020). Observed points marked with an X show values significantly different than expected during pandemic period. Includes 72 months beginning January 2015 and ending December 2020. Expected values were generated from a time series model using observed rates from January 2015 through February 2020. The first 18 months were lost to modeling.

**Figure 3.**
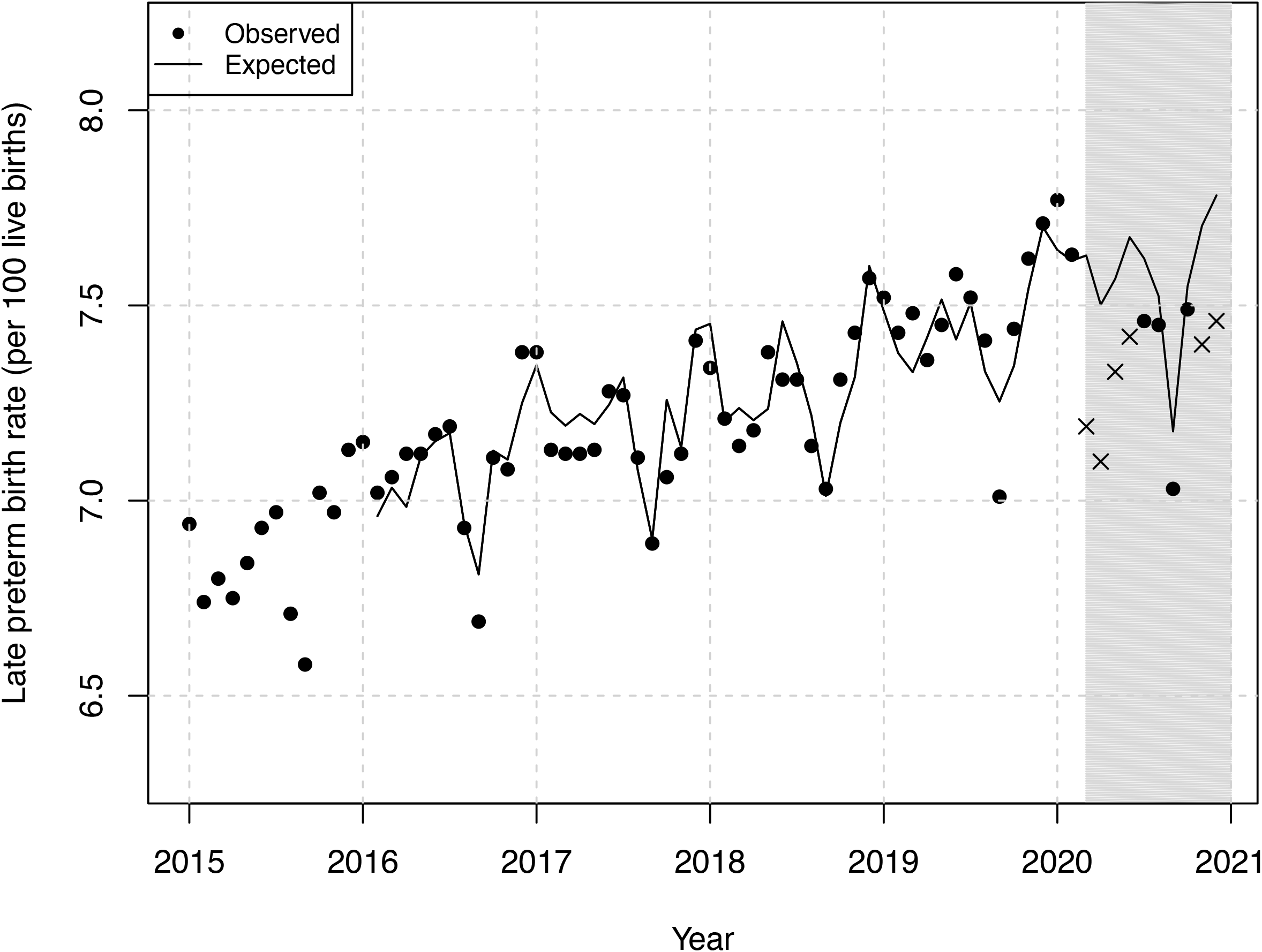
Observed and expected monthly trend of the late preterm birth rate (per 100 live births), 2015-2020. Shaded area represents pandemic months (March through December 2020). Observed points marked with an X show values significantly different than expected during pandemic period. Includes 72 months beginning January 2015 and ending December 2020. Expected values were generated from a time series model using observed rates from January 2015 through February 2020. The first 13 months were lost to modeling.

**Figure 4.**
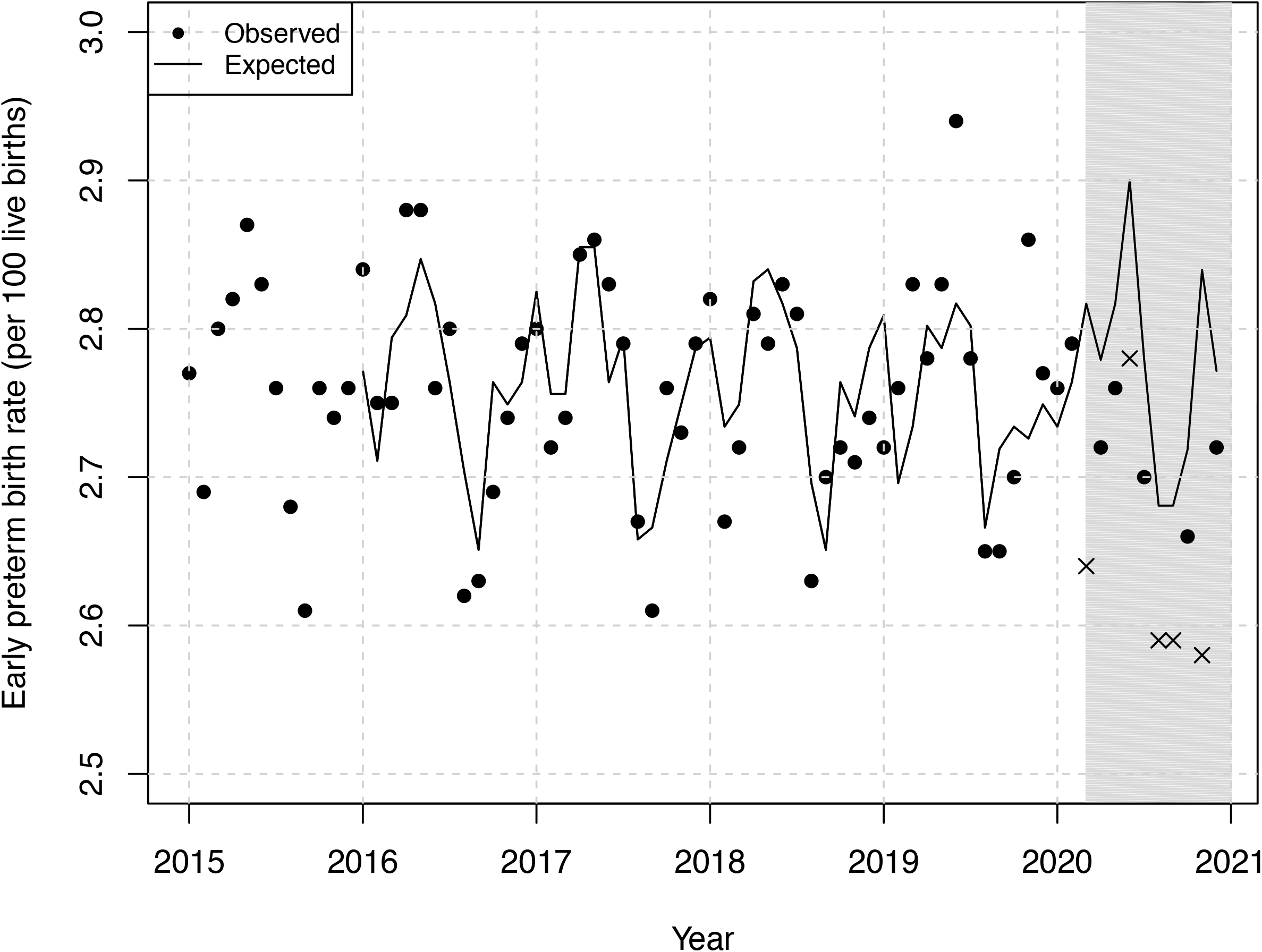
Observed and expected monthly trend of the early preterm birth rate (per 100 live births), 2015-2020. Shaded area represents pandemic months (March through December 2020). Observed points marked with an X show values significantly different than expected during pandemic period. Includes 72 months beginning January 2015 and ending December 2020. Expected values were generated from a time series model using observed rates from January 2015 through February 2020. The first 12 months were lost to modeling.

**Figure 5.**
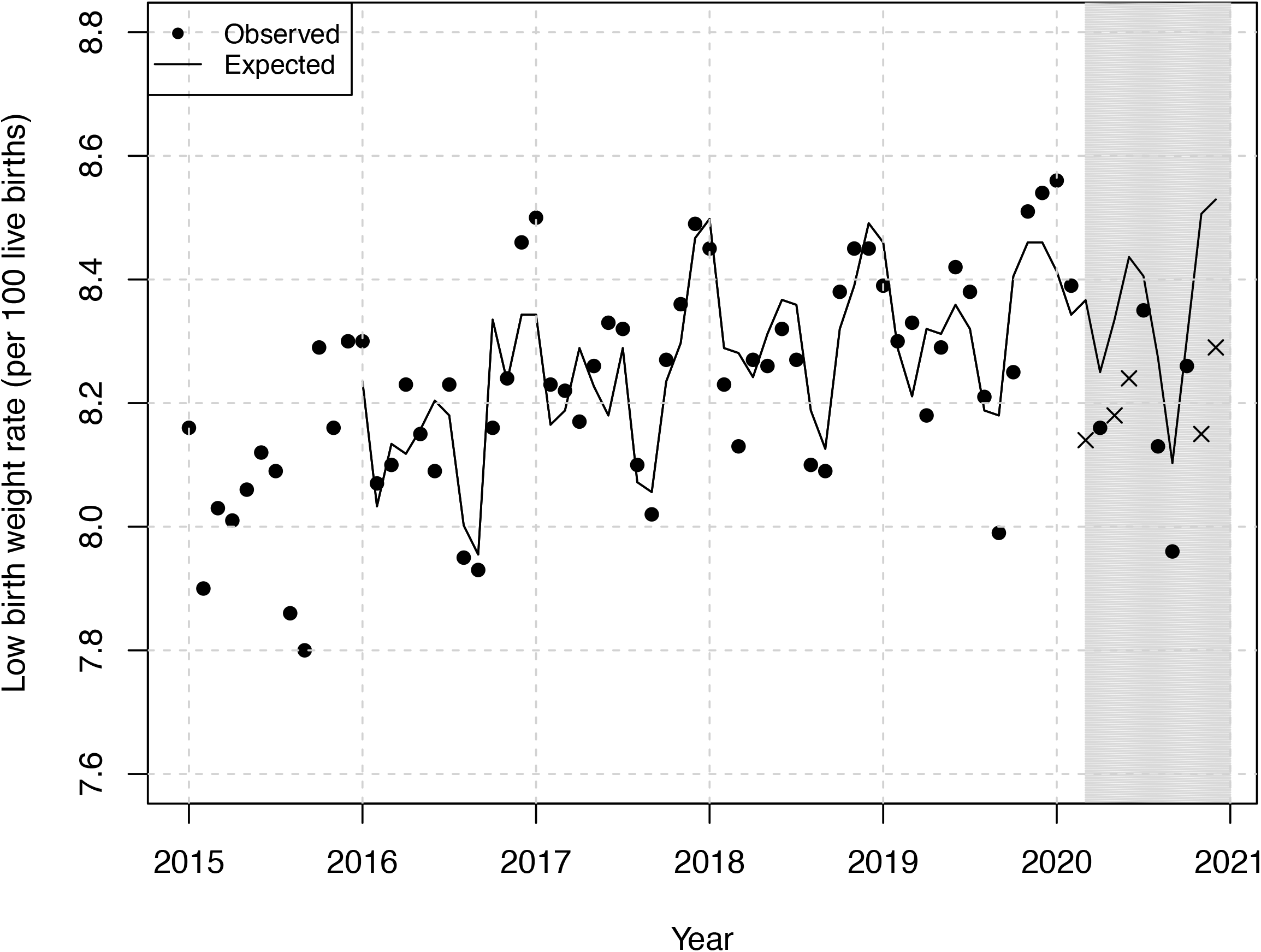
Observed and expected monthly trend of the low birthweight birth rate (per 100 live births), 2015-2020. Shaded area represents pandemic months (March through December 2020). Observed points marked with an X show values significantly different than expected during pandemic period. Includes 72 months beginning January 2015 and ending December 2020. Expected values were generated from a time series model using observed rates from January 2015 through February 2020. The first 12 months were lost to modeling.

**Figure 6.**
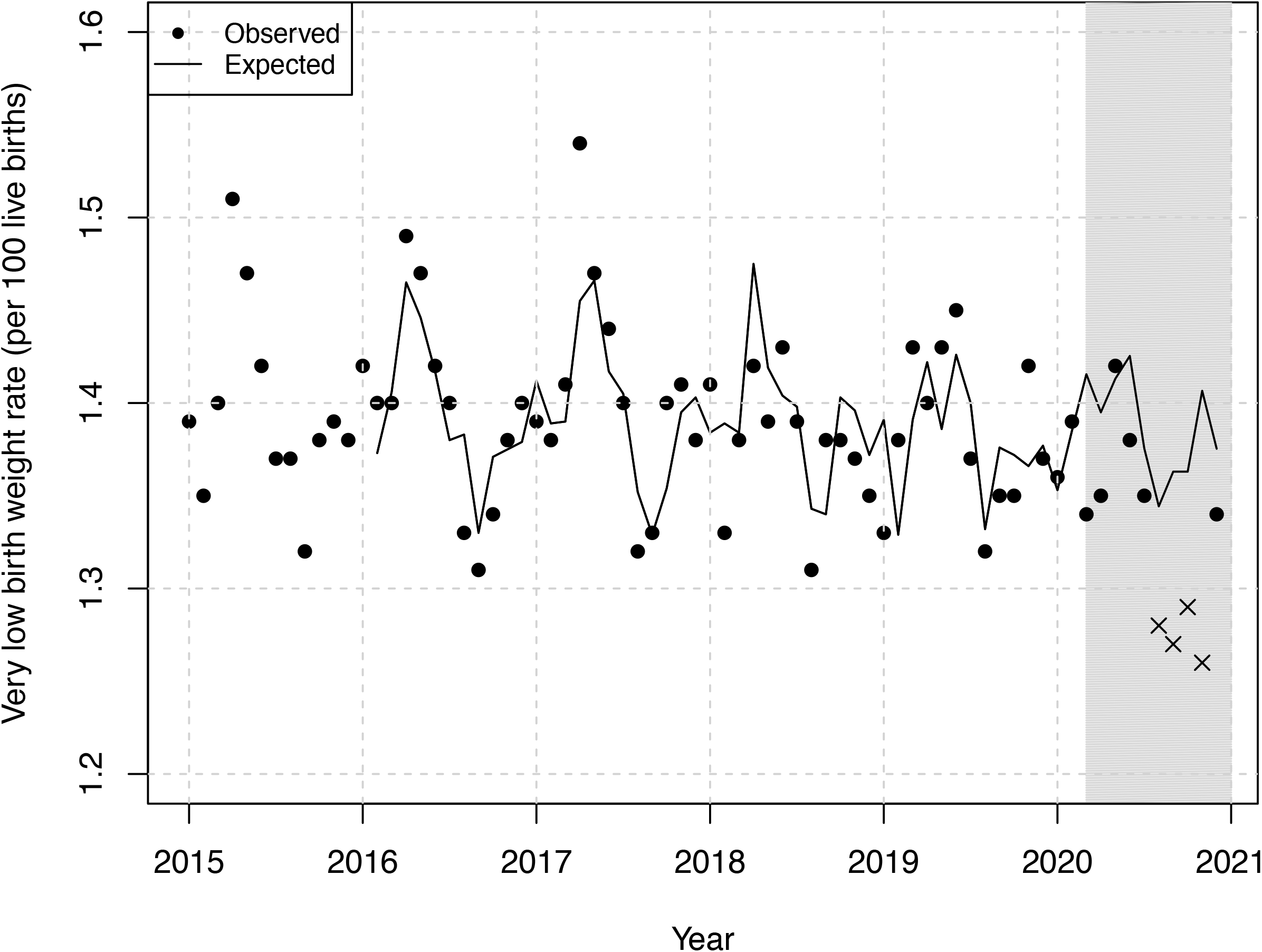
Observed and expected monthly trend of the very low birthweight birth rate (per 100 live births), 2015-2020. Shaded area represents pandemic months (March through December 2020). Observed points marked with an X show values significantly different than expected during pandemic period. Includes 72 months beginning January 2015 and ending December 2020. Expected values were generated from a time series model using observed rates from January 2015 through February 2020. The first 13 months were lost to modeling.

**Figure 7.**
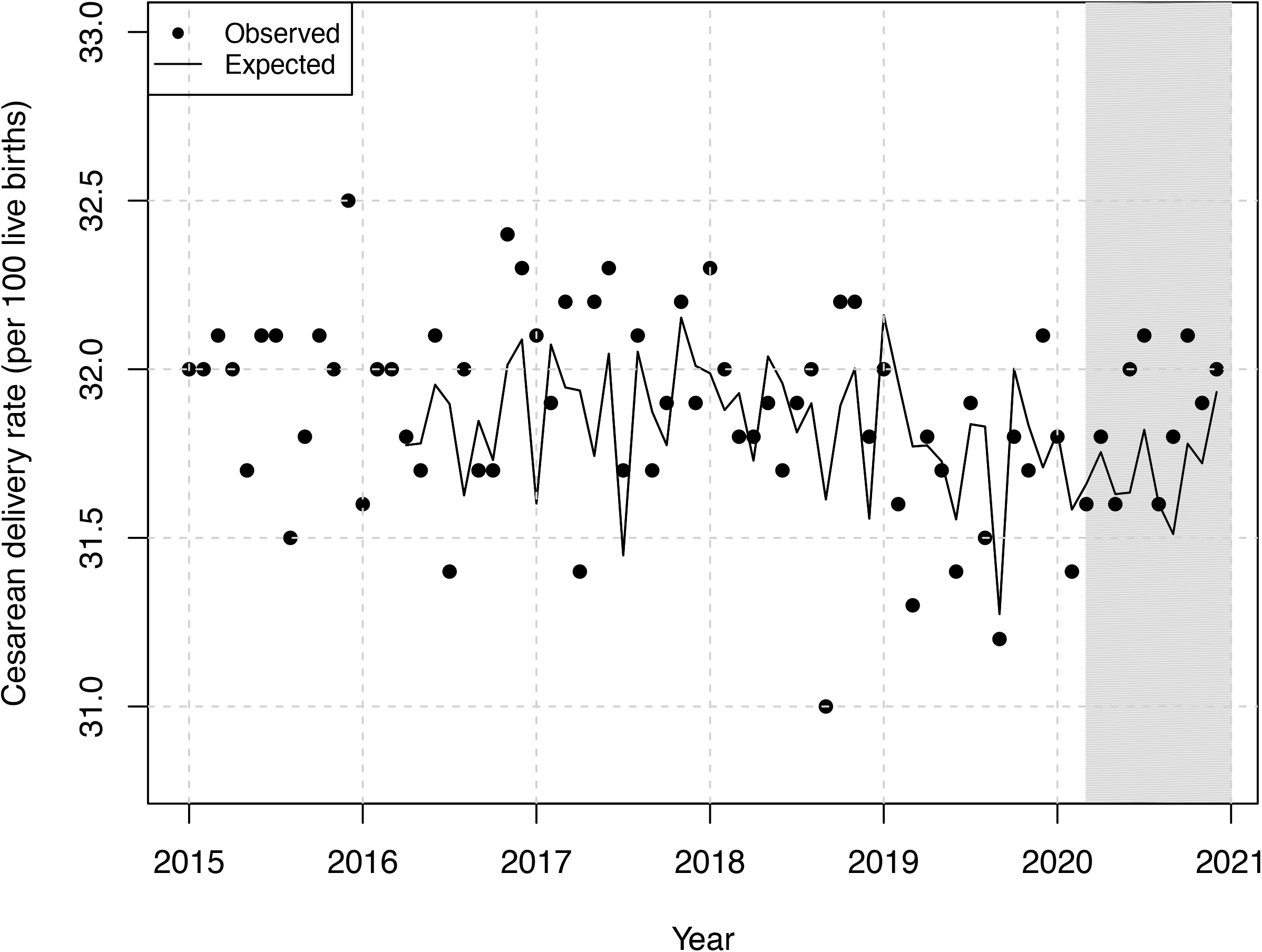
Observed and expected monthly trend of the cesarean delivery rate (per 100 live births), 2015-2020. Shaded area represents pandemic months (March through December 2020). Includes 72 months beginning January 2015 and ending December 2020. Expected values were generated from a time series model using observed rates from January 2015 through February 2020. The first 15 months were lost to modeling.

For total births as well as five of the six indicators (i.e., all but the rate of cesarean delivery), observed values fall well below expected levels (p<.0001 for each test) during the entire pandemic period. Declines in preterm birth and low birth weight were largest in magnitude in both early and later stages of the 2020 pandemic, while those for live births occurred at the end of the year.

## Discussion

Our findings provide some of the first national evidence of substantial reductions in livebirths and adverse perinatal outcomes during the SARS-CoV-2 pandemic. Only cesarean delivery appeared unaffected. These declines were not uniform across the pandemic, suggesting that several mechanisms, which require further study, may explain these patterns.

A key limitation of this study is that the released provisional data from 2020 did not allow us to examine other outcomes, particular subgroups, or severely affected geographies. Future detailed data will allow researchers to study the patterns we identify here in greater detail, including changes in fertility decisions and other proposed mechanisms (e.g., elevated selection *in utero*, reduced air pollution, and changes in clinical practice) that may have caused the observed outcomes.

## Data Availability

All data are publicly available.

https://www.cdc.gov/nchs/covid19/technical-notes-outcomes.htm

http://wonder.cdc.gov/natality.html

## Supplement

### sMethods. Data analysis

**sTable 1**. Time-series results predicting births and perinatal outcomes in the US during the 10 months in 2020 during the SARS-CoV-2 pandemic.

**sTable 2**. Observed and expected monthly values of live births (counts) and perinatal indicators (rates per 100 live births), March through December, 2020.

### sMethods. Data analysis

Our approach follows in the tradition of the interrupted time-series quasi-experiment.^1^ We examine whether the count of live births in the US, and the incidence of several perinatal outcomes (e.g., preterm birth), differ from expected values during the SARS-CoV-2 pandemic beginning March 2020. Births and perinatal outcomes, however, show well-documented patterns such as seasonality, trend, and the tendency for high or low values to be “remembered” into subsequent months.^2^ These patterns, referred to collectively as autocorrelation, violate the assumption of tests of association because the expected value of a patterned series is not its mean. To address this issue, researchers have devised data-driven routines that identify and remove patterns in the outcome variable. We used the routines devised by Box and Jenkins to implement this approach.^3^ These routines express autocorrelation as a product of “autoregressive” (AR), “integrated” (I) and “moving average” (MA) parameters, collectively referred to as ARIMA models. The residuals of these ARIMA models meet the assumptions of correlational tests in that the expected value is zero and the monthly observations are statistically independent of one another.

For each outcome variable, we proceeded through six steps. First, we estimated initial models for the pre-COVID series (62 months beginning January 2015 and ending February 2020) using software from Scientific Computing Associates (version 5.4.6, SCA Corp., Villa Park, IL). Second, we inspected the error term for autocorrelation and, if needed, inserted Box-Jenkins ARIMA parameters to remove any temporal patterns. Third, we added the binary independent variable (i.e., SARS-CoV-2 pandemic) to the best-fitting Box-Jenkins models of each outcome and re-estimated the time-series equation for all 72 months from January 2015 to December 2020. We inspected a concurrent response for the initial spread and shutdown measures (March 2020). We also specified lags of up to 10 months (i.e., from April 2020 to December 2020) given that (i.) the SARS-CoV-2 pandemic persisted through December 2020, and (ii.) fertility decisions in response to the SARS-CoV-2 pandemic may manifest as changes in birth counts and perinatal rates 9 or 10 months later. Fourth, we inspected the residual values of the error term to ensure that they exhibited no temporal patterns. Results, shown in eTable 1, indicate that all outcome series required removal of seasonality, and all but one series (i.e., early preterm) required removal of additional autocorrelation via the inclusion of non-seasonal AR error terms.

1. Bernal JL, Cummins S, Gasparrini A. Interrupted time series regression for the evaluation of public health interventions: a tutorial. *Int J Epidemiol* 2017; 46(1): 348–355.
2. Darrow LA, Strickland MJ, Klein M, et al. Seasonality of birth and implications for temporal studies of preterm birth. *Epidemiology* 2009;20(5):699-706.
3. Box G, Jenkins G, Reinsel G. *Time Series Analysis: Forecasting and Control*. London: Prentice Hall, 1994.

**sTable 1.** Time-series results predicting births and birth outcomes in the US during the 10 months in 2020 during the SARS-CoV-2 pandemic as a function of autocorrelation in n=72 months, from Jan 2015 to Dec 2020. Coefficients represent counts (for births) or risk differences (for all other rates). 95% confidence intervals appear in parentheses.

|  | **Births** | **Preterm** | **Early Preterm** | **Late Preterm** |
| --- | --- | --- | --- | --- |
| Constant | 198151.45 (-4227.35, 400580.25) | 10.84 (9.93, 11.74) | 2.77 (2.72, 2.82) | 8.63 (5.91, 11.35) |
| COVID |  |  |  |  |
| Mar '20 | 1598.48 (-6791.38, 9988.34) | -0.64 (-0.85, -0.44) | -0.17 (-0.26, -0.08) | -0.44 (-0.61, -0.26) |
| Apr '20 | -4239.91 (-12680.89, 4201.06) | -0.56 (-0.79, -0.33) | -0.06 (-0.15, 0.03) | -0.40 (-0.58, -0.21) |
| May '20 | -10407.15 (-18862.71, -1951.58) | -0.34 (-0.56, -0.11) | -0.05 (-0.14, 0.04) | -0.24 (-0.43, -0.05) |
| Jun '20 | 2405.74 (-5993.28, 10804.76) | -0.44 (-0.68, -0.2) | -0.11 (-0.21, -0.01) | -0.26 (-0.46, -0.06) |
| Jul '20 | -6530.90 (-14897.31, 1835.52) | -0.29 (-0.51, -0.06) | -0.08 (-0.17, 0.01) | -0.17 (-0.36, 0.03) |
| Aug '20 | -16202.41 (-24686.35, -7718.48) | -0.19 (-0.43, 0.05) | -0.10 (-0.19, 0.00) | -0.08 (-0.27, 0.12) |
| Sep '20 | -9041.75 (-17354.77, -728.73) | -0.30 (-0.56, -0.04) | -0.10 (-0.19, 0.00) | -0.13 (-0.32, 0.06) |
| Oct '20 | -15098.00 (-23407.97, -6788.02) | -0.13 (-0.38, 0.11) | -0.06 (-0.15, 0.03) | -0.06 (-0.26, 0.13) |
| Nov '20 | -12189.40 (-20675.43, -3703.37) | -0.59 (-0.85, -0.34) | -0.25 (-0.35, -0.16) | -0.32 (-0.52, -0.11) |
| Dec '20 | -18879.34 (-27231.23, -10527.44) | -0.38 (-0.63, -0.13) | -0.05 (-0.14, 0.04) | -0.34 (-0.55, -0.13) |
| AR at lags: | 3, 12 | 1, 5, 12 | 12 | 1, 12 |
| MA at lags: | -- | -- | -- | -- |

|  | **Low birth weight** | **Very low birth weight** | **Cesarean Delivery** |
| --- | --- | --- | --- |
| Constant | 8.5 (8.26, 8.73) | 1.38 (1.36, 1.41) | 31.75 (31.53, 31.96) |
| COVID |  |  |  |
| Mar '20 | -0.24 (-0.39, -0.1) | -0.06 (-0.12, 0.00) | -0.05 (-0.55, 0.44) |
| Apr '20 | -0.10 (-0.25, 0.05) | -0.04 (-0.1, 0.02) | 0.05 (-0.41, 0.51) |
| May '20 | -0.16 (-0.31, -0.01) | 0.01 (-0.05, 0.07) | -0.03 (-0.5, 0.44) |
| Jun '20 | -0.20 (-0.35, -0.04) | -0.04 (-0.1, 0.02) | 0.37 (-0.14, 0.89) |
| Jul '20 | -0.06 (-0.21, 0.1) | -0.03 (-0.09, 0.03) | 0.29 (-0.2, 0.78) |
| Aug '20 | -0.15 (-0.3, 0) | -0.07 (-0.13, 0) | 0 (-0.5, 0.5) |
| Sep '20 | -0.15 (-0.31, 0.01) | -0.1 (-0.16, -0.03) | 0.29 (-0.24, 0.82) |
| Oct '20 | -0.05 (-0.2, 0.1) | -0.08 (-0.14, -0.01) | 0.33 (-0.16, 0.83) |
| Nov '20 | -0.36 (-0.52, -0.2) | -0.14 (-0.21, -0.08) | 0.19 (-0.31, 0.68) |
| Dec '20 | -0.24 (-0.4, -0.08) | -0.04 (-0.1, 0.03) | 0.09 (-0.41, 0.59) |
| AR at lags | 1, 12 | 1, 4, 12 | 3, 12 |
| MA at lags: | -- | -- | -- |

**sTable 2.**
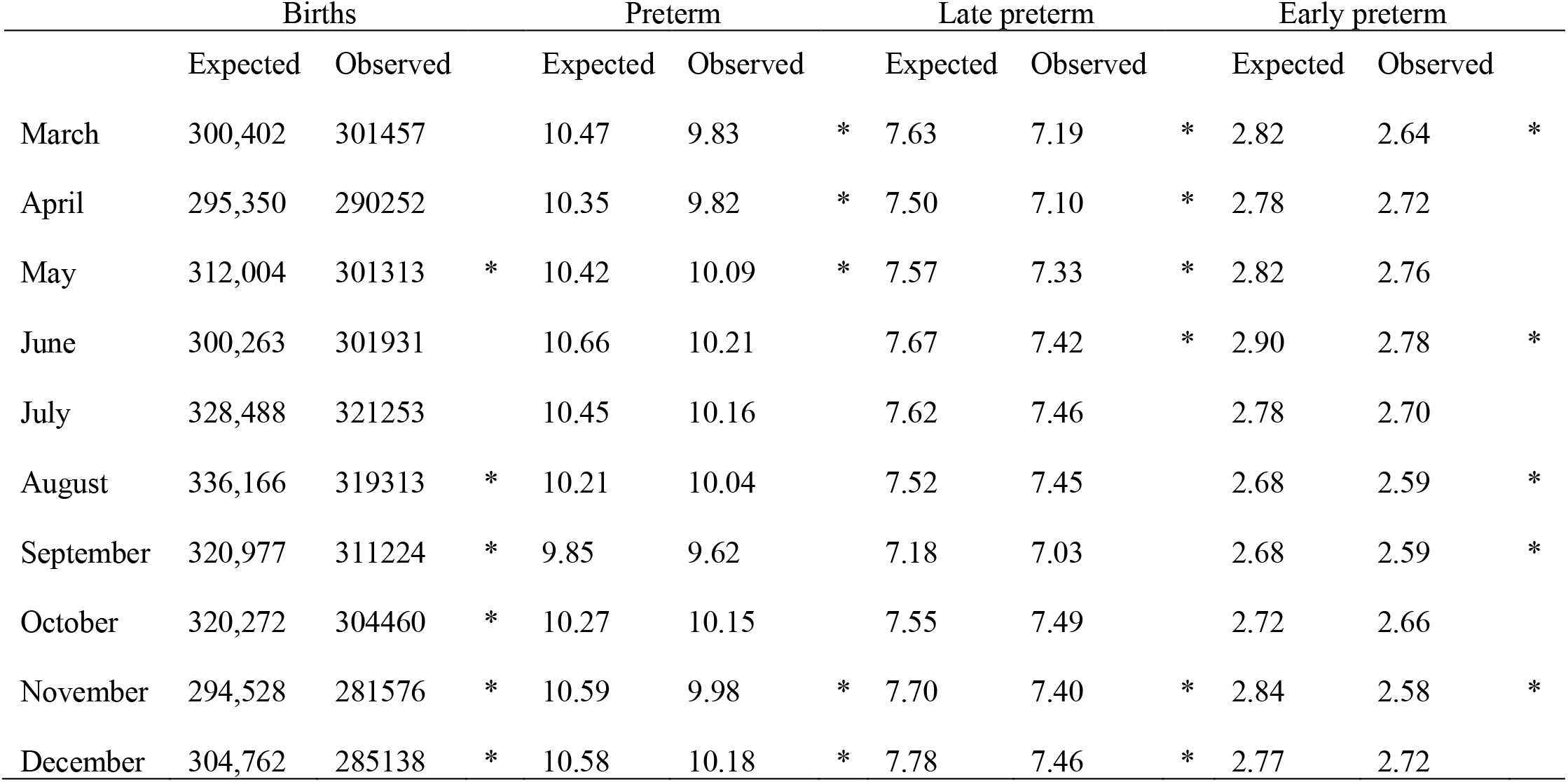
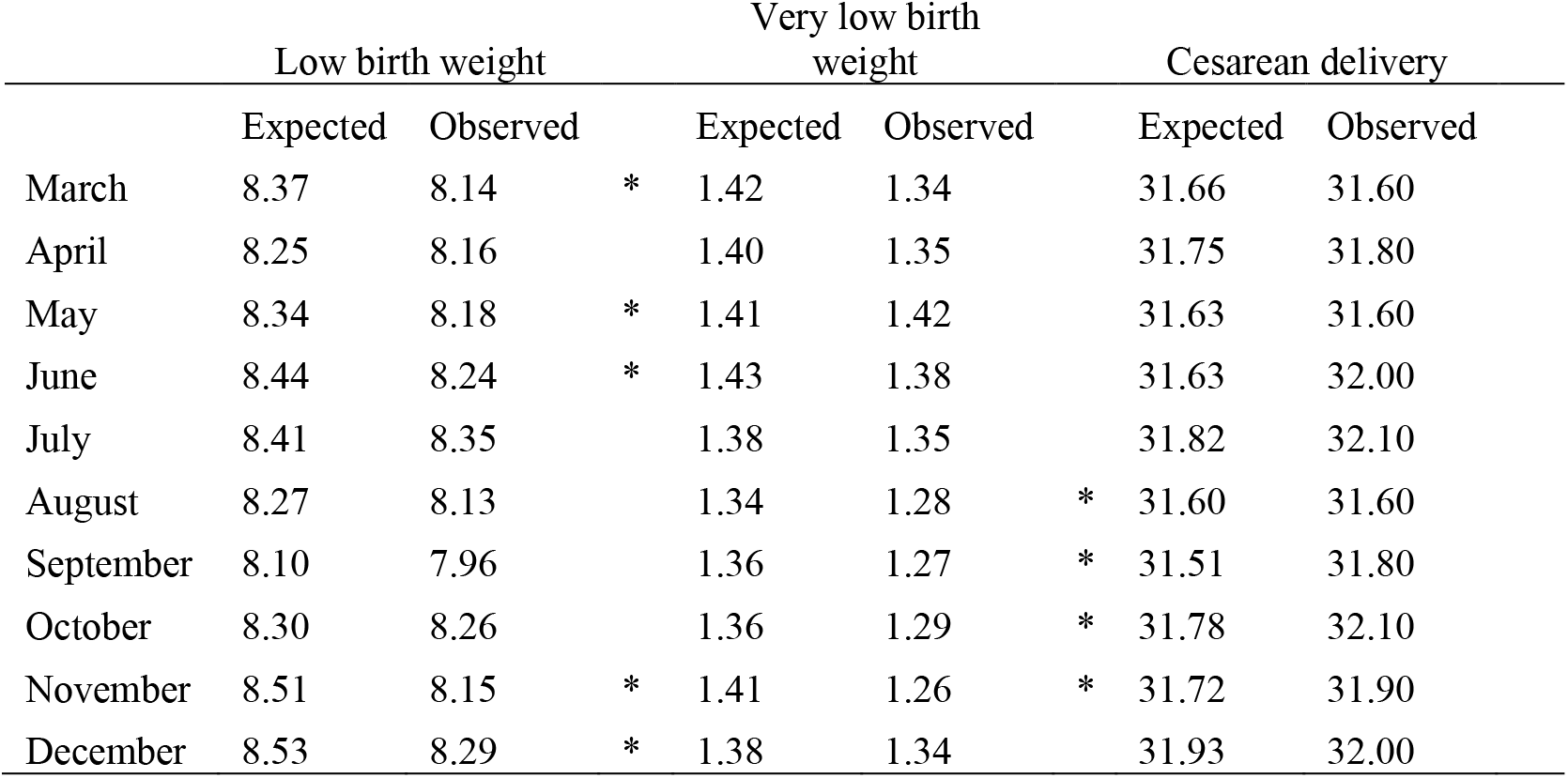
Observed and expected monthly values of live births (counts) and perinatal indicators (rates per 100 live births), March through December, 2020.

